# Bacillus Calmette-Guérin scar reactivation (BCGitis) in Kawasaki disease. A Scoping Review

**DOI:** 10.1101/2025.10.03.25337219

**Authors:** Hassib Narchi, Alyson Skinner

**Author notes:** Corresponding Author: Hassib Narchi, Department of Paediatrics, College of Medicine & Health Sciences, United Arab Emirates University, PO Box 15551, Al Ain, United Arab Emirates; ☏.

## Abstract

**Aim:** To map and synthesise the evidence on the scattered and heterogeneous reported prevalence of Bacillus Calmette-Guérin scar reactivation (BCGitis) in Kawasaki disease (KD), examine clinical associations, and identify knowledge gaps for future research.

**Methods:** A scoping review was conducted in accordance with PRISMA-ScR guidelines. PubMed, Scopus, EMBASE, Web of Science, and Cochrane Library were searched for observational studies of BCG-vaccinated children with KD. We retrieved the prevalence of BCGitis and its associations with age, sex, KD form, BCG vaccine strain, coronary artery abnormalities (CAA), and treatment response.

**Results:** Fifteen studies from five Asian countries (Japan, Korea, Taiwan, China, Singapore), including 21,880 children with KD, were identified. The reported prevalence of BCGitis ranged from 3.5% to 75.2% (overall prevalence 40.1%, 95% CI 39.6–40.6), with a higher prevalence in younger children. It was associated with lower CAA rates (11.8% vs 13.8%) but showed no significant association with KD type, vaccine strain, or treatment response.

**Conclusion:** BCGitis is a common manifestation of KD in BCG-vaccinated children and may have diagnostic and prognostic value, particularly in younger children and those with incomplete presentations.

## 1. INTRODUCTION

Early diagnosis and treatment of Kawasaki disease (KD) are essential for preventing the development of coronary artery abnormalities (CAA).^1,2^ Diagnosis remains primarily clinical.^3,4^ Incomplete presentations, difficult to diagnose, are more prevalent in infants in whom CAA are more common.^2,4–8^ The absence of pathognomonic biomarkers necessitates the inclusion of other clinical indicators to facilitate timely diagnosis and treatment.

Scar reactivation at the Bacillus Calmette-Guérin (BCG) vaccination site is also known as “BCGitis”. It may occur as an early sign of KD in countries with universal BCG vaccination programs. ^5,8–11^ It has been included in the Japanese KD diagnostic guidelines in 2019 and is also used in Korea. ^6,12,13^ There is heterogeneity of the BCGitis prevalence rates in the reported studies, ranging from < 5% to > 70%. This reflects differences in research methodologies, healthcare systems, population characteristics, BCG vaccination programs, vaccine strains used, and inconsistent clinical assessment for BCGitis.

Evidence-based diagnostic standards and clinical decision-making algorithms depend on precise prevalence data, identifying clinical factors and responses to treatment associated with BCGitis. This could improve risk stratification and targeted assessment strategies. Consolidating all such information in a “one-stop” resource is the aim of this scoping review. The intent is to map the extent of available evidence, highlight knowledge gaps and guide future research.

## 2. METHODS

This scoping review followed the guidelines for Preferred Reporting Items for Systematic Reviews and Meta-Analyses extension for Scoping Reviews (PRISMA-ScR) (Supplementary File 6).

### 2.1 Information Sources

Five electronic databases were searched: PubMed, Scopus, EMBASE, Web of Science Core Collection, and Cochrane Library. Reference lists in retrieved studies were hand-searched. Grey literature was not included, as the focus was on peer-reviewed evidence.

### 2.2 Search Strategy

The search was based on the Population, Concept, Context (PCC) framework. The Population consisted of children (<18 years) with KD. The Concept was: BCG scar reactivation/ erythema/ induration/ suppuration (“BCGitis”). The Context was: any clinical setting or country. Search strategies are provided in Supplementary File 4.

### 2.3 Inclusion Criteria

- Original observational studies (cohort, cross-sectional, case-control)
- Children (<18 years) with KD, hospital- or clinic-based
- BCG-vaccinated children, or studies providing separate data for vaccinated subgroups
- Systematic assessment of BCG vaccination sites
- Published in English, full-text available, without date or geographic restrictions

### 2.4 Exclusion Criteria

- Case reports, case series, reviews, editorials, or commentaries
- Overlap of patients among studies
- Unknown or unverified BCG vaccination status
- Incidental finding of BCGitis without systematic assessment
- Animal or experimental studies

### 2.5 Study Selection

Two reviewers (HN and AS) independently screened titles and abstracts. Duplicates were removed, and potentially eligible studies underwent full-text review. Disagreements were resolved by consensus. Reasons for exclusion were recorded.

### 2.6 Data Extraction

Data extraction was performed independently by two reviewers using a standardised form (Supplementary File 5). Discrepancies were resolved by discussion and re-checking the original reports. Authors were not contacted for missing data, consistent with scoping review methodology

### 2.7 Data Charting

Subgroup analyses were conducted for age groups, sex, BCG vaccination strain, KD form, and CAA development. We also compared BCGitis prevalence with other KD diagnostic features, treatment response, and analysed its timing in the course of KD.

### 2.8 Synthesis of Results

Consistent with scoping review methodology, data synthesis and results were primarily descriptive and narrative rather than statistical and meta-analytical. Prevalence estimates were calculated where appropriate.

### 2.9 Quality Assessment

Formal quality assessment or evaluation of risk of bias analysis are not required for a scoping review.^14^

## 3. RESULTS

### 3.1 Description of Included Studies

The search identified 577 records (PubMed: 204, Scopus: 323, EMBASE: 23, Web of Science: 6, Cochrane Library: 21). After removing 432 duplicates, 145 studies were screened by title and abstract. Of these, 75 were excluded as irrelevant. Full-text review was performed for 70 studies, of which 55 were excluded: no systematic BCGitis assessment (n=11), unknown vaccination status (n=6), case reports (n=13), reviews (n=7), overlapping reported populations (n=1), non-English language (n=3), conference abstracts (n=2), and editorials/commentaries/letters (n=4). Fifteen studies met all criteria and were included. The selection process is shown in the PRISMA flow diagram (Fig. 1). No additional studies were identified from reference lists.

**Figure 1.**
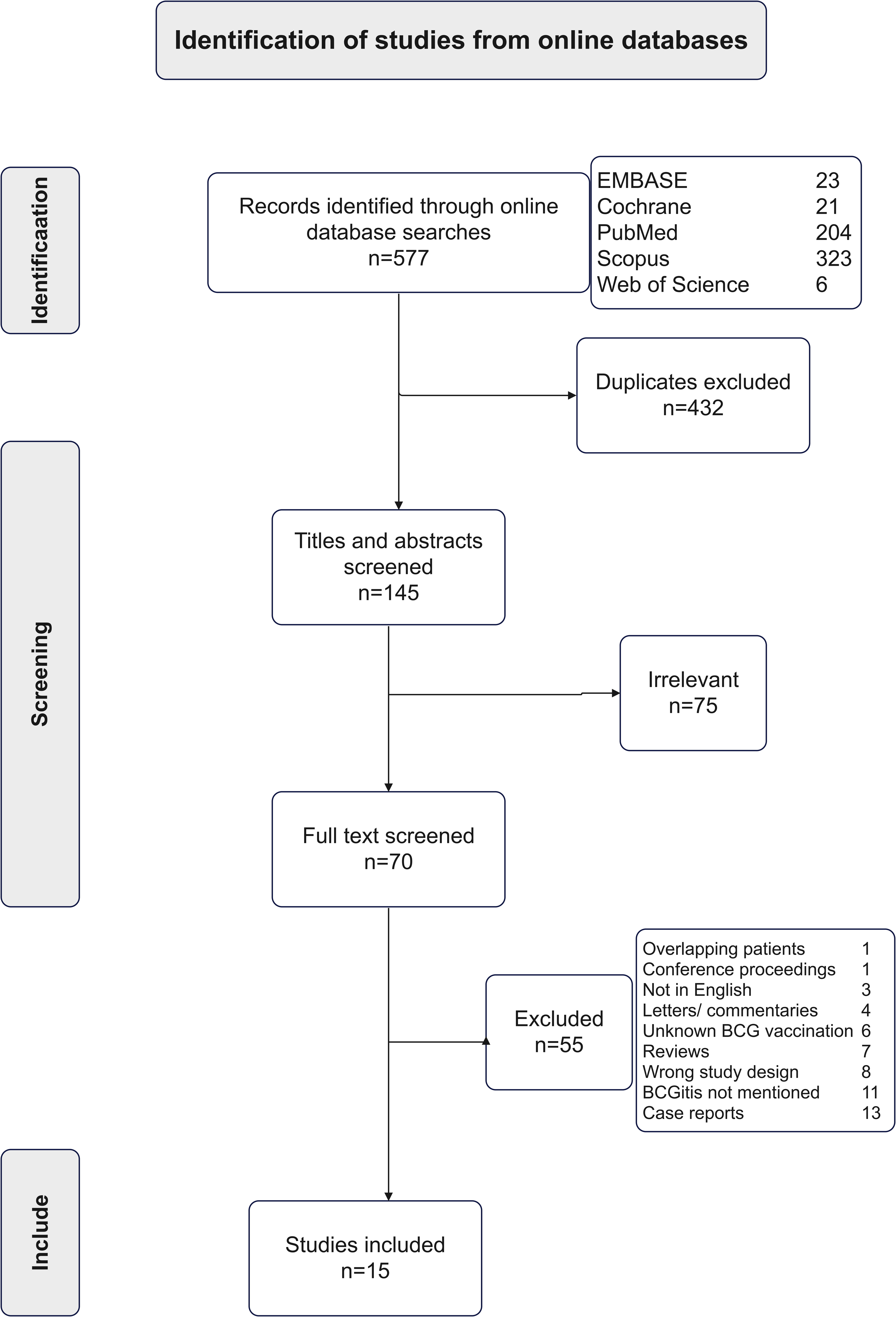
Flow diagram of selected studies.

The 15 studies were published between 1993 and 2023. They included 21,880 children with KD, of whom 8,781 (40%) had BCGitis. They originated from Japan (n=3), Korea (n=5), Taiwan (n=4), China (n=2), and Singapore (n=1). Fourteen studies (93%) were published after 2010, reflecting increasing recognition of BCGitis. Twelve were retrospective observational studies, two were case-control, and one was a clinical immunology study. All were hospital- or clinic-based (Tables 1 and 2).. All participants were BCG-vaccinated as per national policy or with vaccination status verified individually, though documentation methods varied.

**Table 1.** Characteristics of 15 included studies of 211,880 children with KD.

| Characteristics |  | n (%) | References |
| --- | --- | --- | --- |
| **Publication year** | <2010 | 1 (6.6) | Yokota<sup>25</sup> |
|  | 2011-2015 | 7 (46.6) | Lin<sup>11</sup>, Seo<sup>6</sup>, Lai<sup>9</sup>, Teng<sup>26</sup>, Kang<sup>27</sup>, Bae<sup>28</sup>, Ruan<sup>24</sup> |
|  | 2016-2020 | 4 (26.6) | Park<sup>5</sup>, Araki<sup>29</sup>, Kim<sup>30</sup>, Loh<sup>1</sup> |
|  | >2021 | 3 (20) | Takikawa<sup>31</sup>, Peng<sup>23</sup>, Yang<sup>20</sup> |
| **Study setting** | Health care facility (hospital, clinic) | 15 (100) | Yokota<sup>25</sup>, Lin<sup>11</sup>, Seo<sup>6</sup>, Lai<sup>9</sup>, Teng<sup>26</sup>, Kang<sup>27</sup>, Bae<sup>28</sup>, Ruan<sup>24</sup>, Park<sup>5</sup>, Araki<sup>29</sup>, Kim<sup>30</sup>, Loh<sup>1</sup>, Takikawa<sup>31</sup>, Peng<sup>23</sup>, Yang<sup>20</sup> |
| **Country** | Korea | 5 (33.3) | Bae<sup>28</sup>, Kang<sup>27</sup>, Park<sup>5</sup> |
|  | Taiwan | 4 (26.6) | Lin<sup>11</sup>, Teng<sup>26</sup>, Lai<sup>9</sup>, Yang<sup>20</sup> |
|  | Japan | 3 (20) | Yokota<sup>25</sup>, Araki<sup>29</sup>, Takikawa<sup>31</sup> |
|  | China | 2 (13.3) | Ruan<sup>24</sup>, Peng<sup>23</sup> |
|  | Singapore | 1 (6.6) | Loh<sup>1</sup> |
| **Age groups comparison** |  | 8 (53.3) | Loh<sup>1</sup>, Peng<sup>23</sup>, Kang<sup>27</sup>, Teng<sup>26</sup>, Park<sup>5</sup>, Lai<sup>9</sup>, Ruan<sup>24</sup>, Takikawa<sup>31</sup>, Kim<sup>30</sup> |
| **Sex groups comparison** |  | 4 (26.6) | Takikawa<sup>31</sup>, Park<sup>5</sup>, Loh<sup>1</sup>, Lai<sup>9</sup> |
| **Study design** | Observational retrospective | 12 (80) | Seo<sup>6</sup>, Lai<sup>9</sup>, Teng<sup>26</sup>, Kang<sup>27</sup>, Bae<sup>28</sup>, Ruan<sup>24</sup>, Park<sup>5</sup>, Araki<sup>29</sup>, Loh<sup>1</sup>, Takikawa<sup>31</sup>, Peng<sup>23</sup>, Yang<sup>20</sup> |
|  | Case-control genetic association study | 2 (13.3) | Lin<sup>11</sup>, Kim<sup>30</sup> |
|  | Clinical immunology observational | 1 (6.6) | Yokota<sup>25</sup> |
| **Data source** | Patients' records | 15 (100) | Yokota<sup>25</sup>, Lin<sup>11</sup>, Seo<sup>6</sup>, Lai<sup>9</sup>, Teng<sup>26</sup>, Kang<sup>27</sup>, Bae<sup>28</sup>, Ruan<sup>24</sup>, Park<sup>5</sup>, Araki<sup>29</sup>, Kim<sup>30</sup>, Loh<sup>1</sup>, Takikawa<sup>31</sup>, Peng<sup>23</sup>, Yang<sup>20</sup> |
| **Variables included** | BCGitis presence | 15 (100) | Yokota<sup>25</sup>, Lin<sup>11</sup>, Seo<sup>6</sup>, Lai<sup>9</sup>, Teng<sup>26</sup>, Kang<sup>27</sup>, Bae<sup>28</sup>, Ruan<sup>24</sup>, Park<sup>5</sup>, Araki<sup>29</sup>, Kim<sup>30</sup>, Loh<sup>1</sup>, Takikawa<sup>31</sup>, Peng<sup>23</sup>, Yang<sup>20</sup> |
|  | Age groups | 6 (40) | Loh<sup>1</sup>, Peng<sup>23</sup>, Kang<sup>27</sup>, Teng<sup>26</sup>, Park<sup>5</sup>, Ruan<sup>24</sup> |
|  | Sex groups | 4 (26.6) | Takikawa<sup>31</sup>, Park<sup>5</sup>, Loh<sup>1</sup>, Lai<sup>9</sup> |
|  | Prior BCG vaccination | 15 (100) | Yokota<sup>25</sup>, Lin<sup>11</sup>, Seo<sup>6</sup>, Lai<sup>9</sup>, Teng<sup>26</sup>, Kang<sup>27</sup>, Bae<sup>28</sup>, Ruan<sup>24</sup>, Park<sup>5</sup>, Araki<sup>29</sup>, Kim<sup>30</sup>, Loh<sup>1</sup>, Takikawa<sup>31</sup>, Peng<sup>23</sup>, Yang<sup>20</sup> |
|  | Complete, incomplete KD | 4 (26.6) | Loh<sup>1</sup>, Peng<sup>23</sup>, Kang<sup>27</sup>, Teng<sup>26</sup> |
|  | Response to IVIG | 3 (20) | Park<sup>5</sup>, Takikawa<sup>31</sup>, Lai<sup>9</sup> |
|  | CAA | 4 (26.6) | Park<sup>5</sup>, Takikawa<sup>31</sup>, Lai<sup>9</sup>, Araki<sup>29</sup> |
|  | KD signs | 7 (46.6) | Lin<sup>11</sup>, Araki<sup>29</sup>, Kang<sup>27</sup>, Teng<sup>26</sup>, Bae<sup>28</sup>, Peng<sup>23</sup>, Ruan<sup>24</sup> |
| BCG: Bacille Calmette-Guérin; KD: Kawasaki disease; CAA: coronary artery abnormalities: |  |  |  |

**Table 2.**
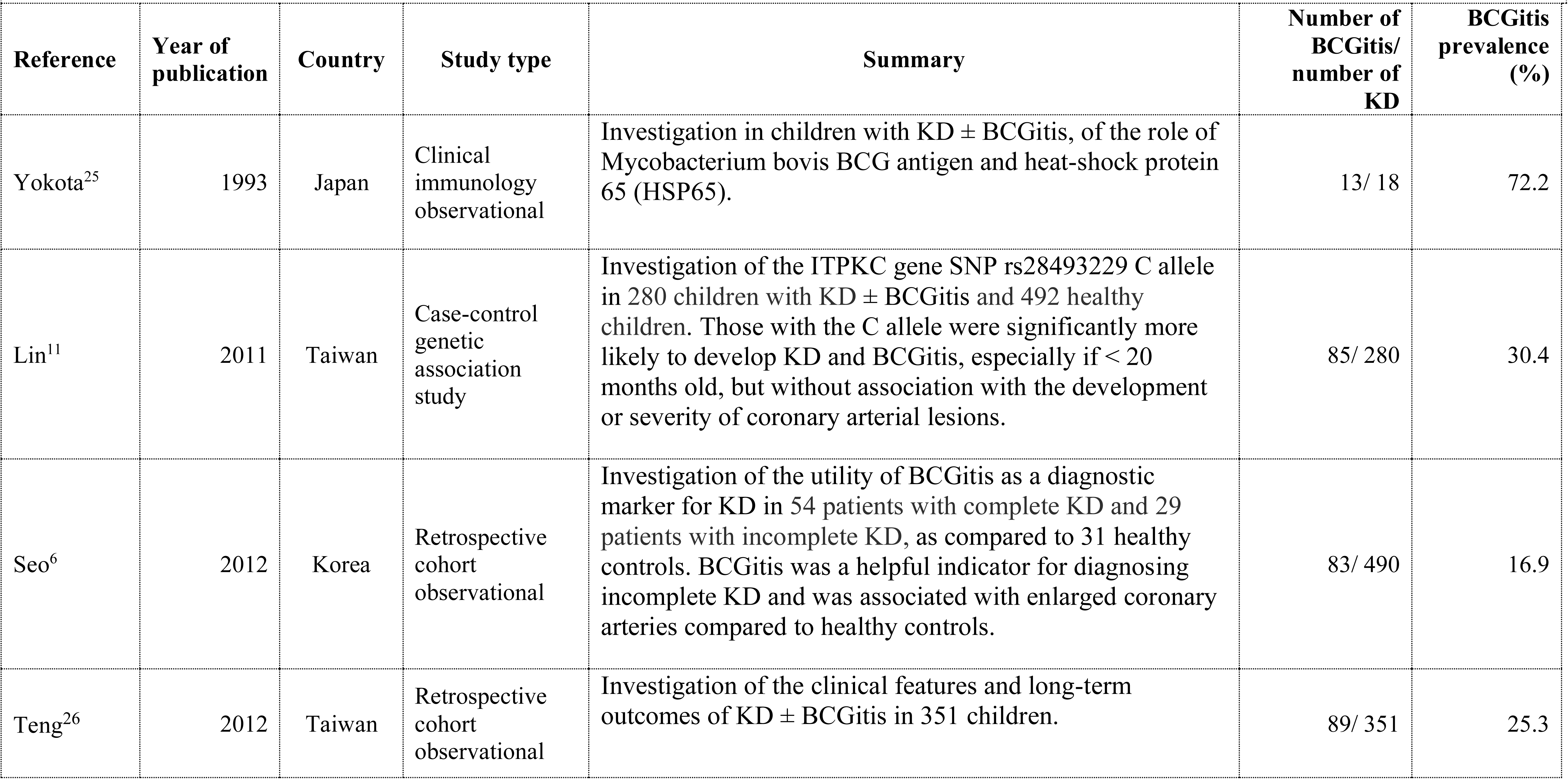
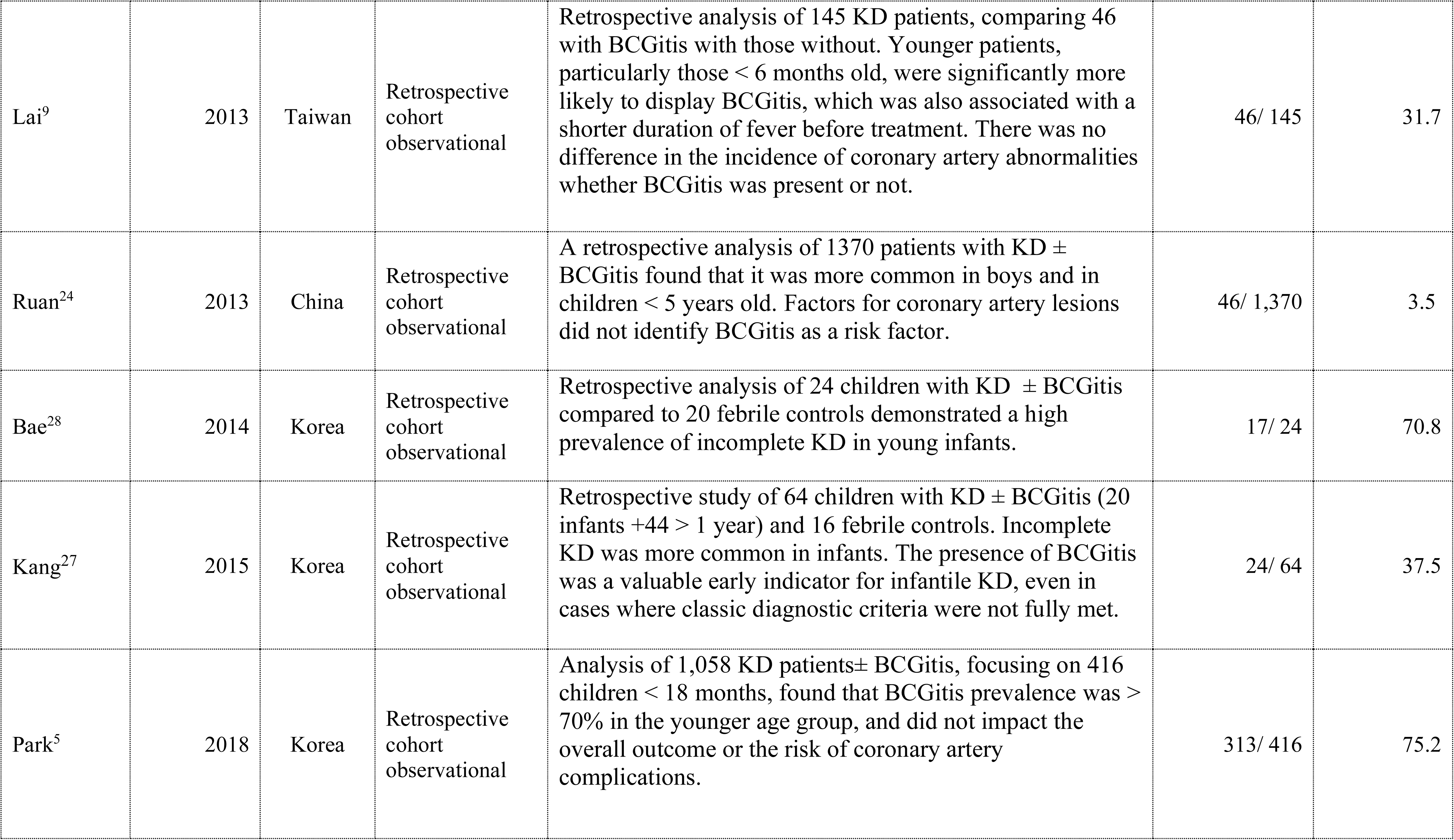
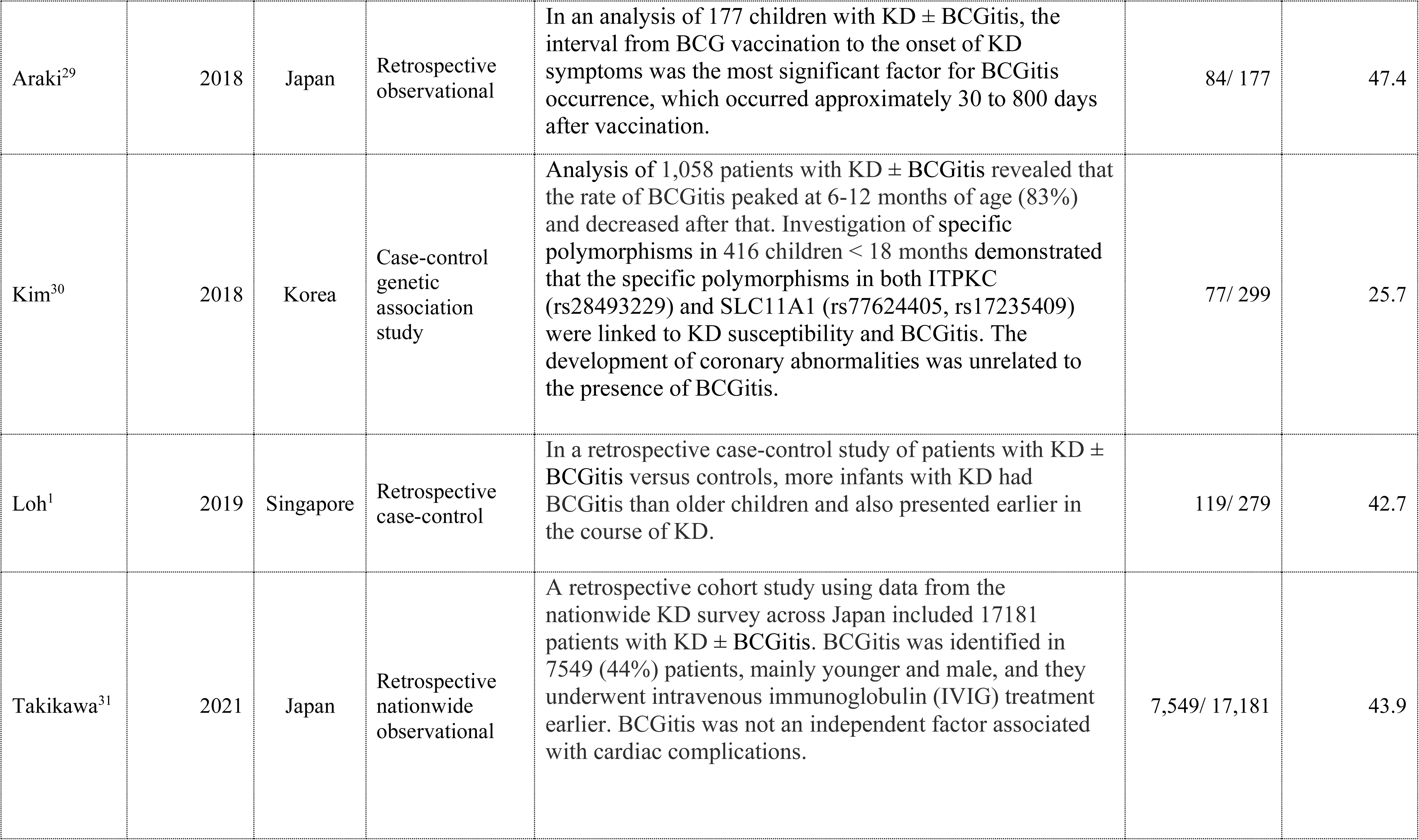
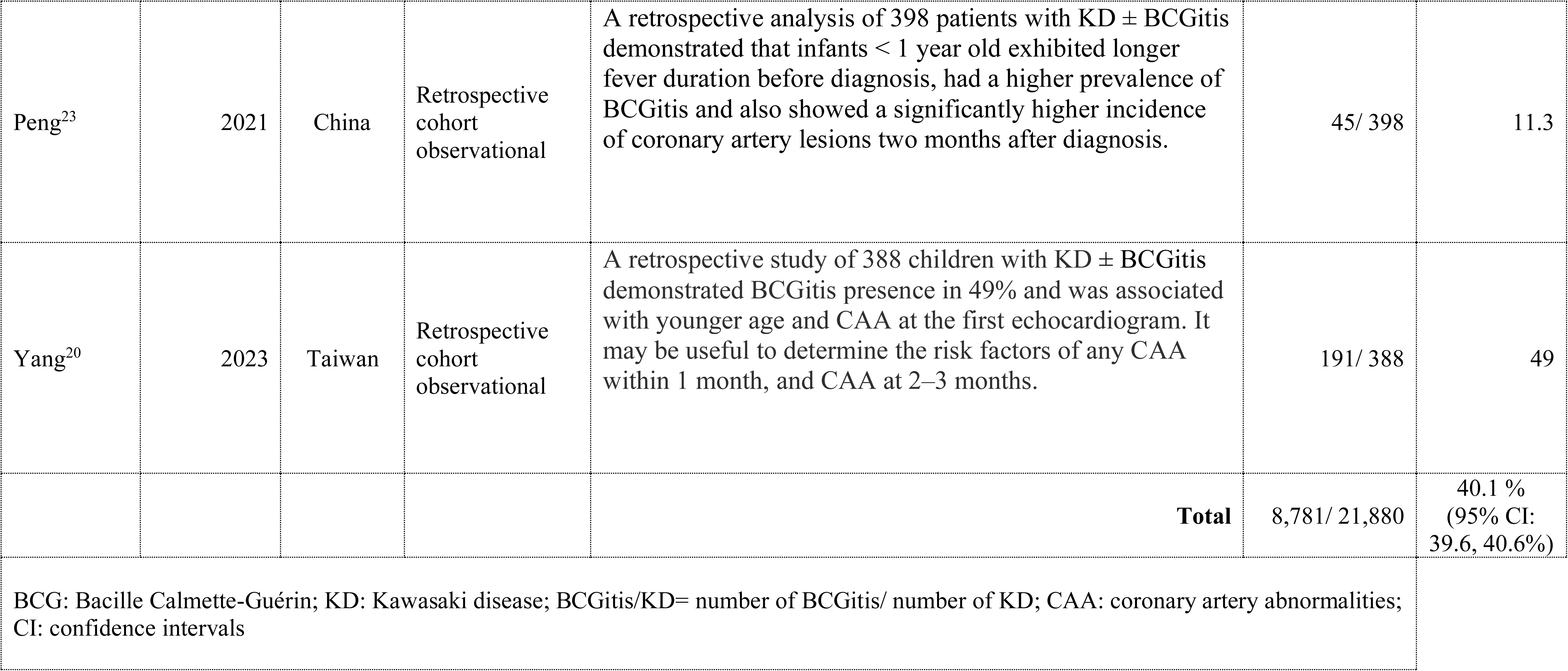
Characteristics of 15 studies, in chronological order, including 21,880 children with KD with reported BCGitis prevalence.

### 3.2 Overall BCGitis prevalence

The prevalence (Table 2) varied from 3.5% (China) to 75.2% (Korea). The overall prevalence was 40.1% (95% confidence intervals 39.6, 40.6). The median prevalence was highest in Japan (43.9%, range 43.9–72.2) and Korea (37.5%, range 16.9–75.2), intermediate in Taiwan (28.5%, range 25.3–49.0), and Singapore 42.7%. It was the lowest in China (7.4%, range 3.5–11.3).

### 3.3 BCGitis prevalence by age

Nine studies (60%) reported age-stratified data. All showed higher prevalence in younger children. Among those <12 months, the prevalence ranged from 18.4% to 80.0%; in children >2 years, it ranged from 0% to 63.0%. Two studies found no BCGitis in children >5 years. The largest analysis by Takikawa (n=17,181) showed a prevalence of 61.8% in children <2 years versus 9.1% in ≥2 years (Supplementary file: Table 1A).

### 3.4 BCGitis prevalence by sex

Four studies (27%) reported sex-specific data, two of which three showed slightly higher rates in males. The overall prevalence was 46.4% in males versus 41.9% in females (Supplementary file: Table 1B).

### 3.5 Prevalence of BCGitis and KD signs between KD forms

#### 3.5.1 Complete vs incomplete Kawasaki disease

Three studies compared the prevalence of BCGitis between KD forms. Two reported a higher prevalence in complete KD (44.1% vs 43.0%; 31.8% vs 13.0%), while one found a higher prevalence in incomplete KD (87.2% vs 71.7%). The overall prevalence was nearly identical between the two forms (44.6% vs 44.3%) (Table 3A).

**Table 3.**
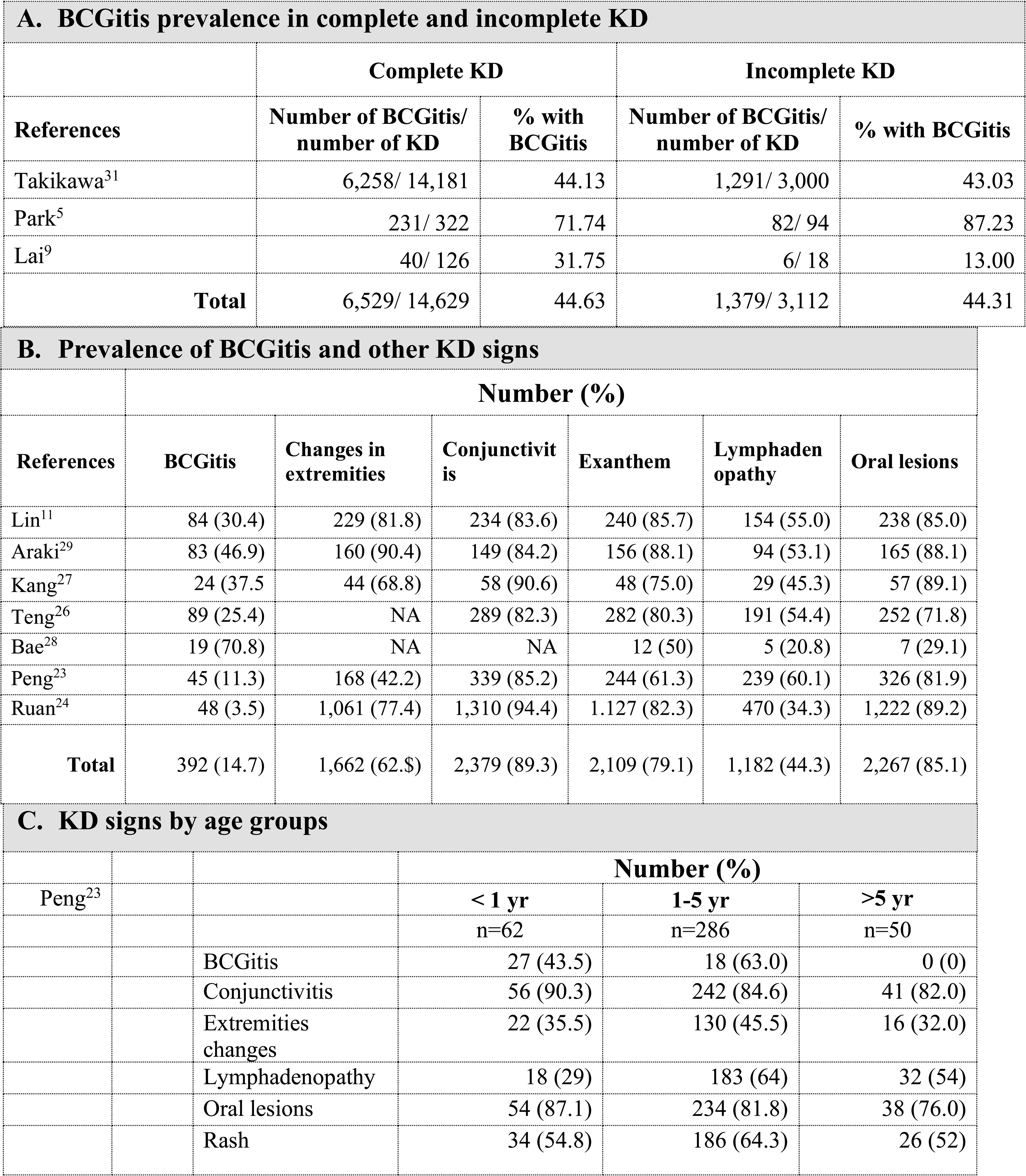
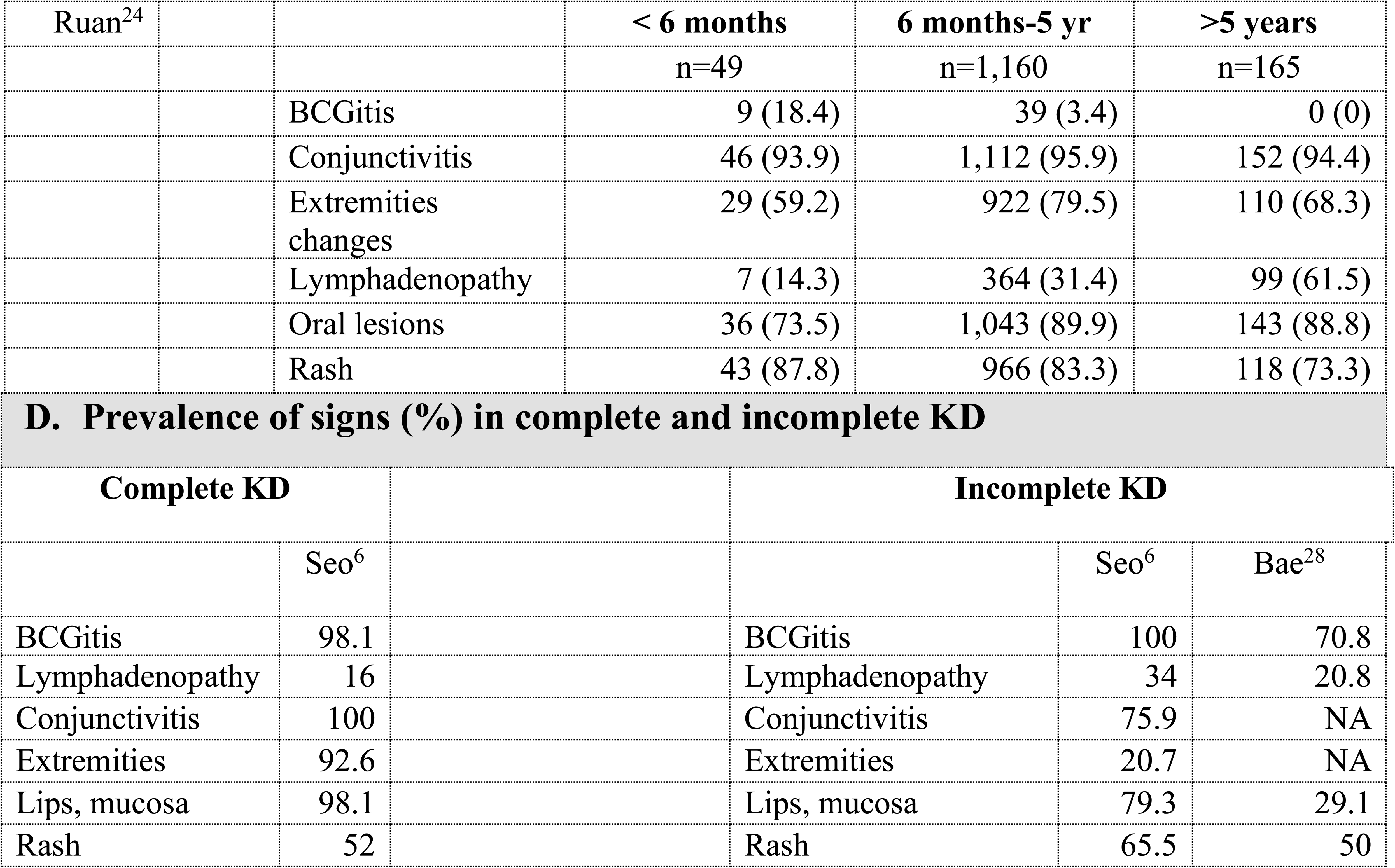
Prevalence of BCGitis and KD clinical manifestations by KD forms.

| A. BCGitis prevalence in complete and incomplete KD |  |  |  |  |  |
| --- | --- | --- | --- | --- | --- |
|  |  | Complete KD |  | Incomplete KD |  |
| References |  | Number of BCGitis/<br>number of KD | % with<br>BCGitis | Number of BCGitis/<br>number of KD | % with BCGitis |
| Takikawa<sup>31</sup> |  | 6,258/ 14,181 | 44.13 | 1,291/ 3,000 | 43.03 |
| Park<sup>5</sup> |  | 231/ 322 | 71.74 | 82/ 94 | 87.23 |
| Lai<sup>9</sup> |  | 40/ 126 | 31.75 | 6/ 18 | 13.00 |
| Total |  | 6,529/ 14,629 | 44.63 | 1,379/ 3,112 | 44.31 |

| B. Prevalence of BCGitis and other KD signs |  |  |  |  |  |  |
| --- | --- | --- | --- | --- | --- | --- |
|  | Number (%) |  |  |  |  |  |
| References | BCGitis | Changes in<br>extremities | Conjunctivit<br>is | Exanthem | Lymphaden<br>opathy | Oral lesions |
| Lin<sup>11</sup> | 84 (30.4) | 229 (81.8) | 234 (83.6) | 240 (85.7) | 154 (55.0) | 238 (85.0) |
| Araki<sup>29</sup> | 83 (46.9) | 160 (90.4) | 149 (84.2) | 156 (88.1) | 94 (53.1) | 165 (88.1) |
| Kang<sup>27</sup> | 24 (37.5 | 44 (68.8) | 58 (90.6) | 48 (75.0) | 29 (45.3) | 57 (89.1) |
| Teng<sup>26</sup> | 89 (25.4) | NA | 289 (82.3) | 282 (80.3) | 191 (54.4) | 252 (71.8) |
| Bae<sup>28</sup> | 19 (70.8) | NA | NA | 12 (50) | 5 (20.8) | 7 (29.1) |
| Peng<sup>23</sup> | 45 (11.3) | 168 (42.2) | 339 (85.2) | 244 (61.3) | 239 (60.1) | 326 (81.9) |
| Ruan<sup>24</sup> | 48 (3.5) | 1,061 (77.4) | 1,310 (94.4) | 1,127 (82.3) | 470 (34.3) | 1,222 (89.2) |
| Total | 392 (14.7) | 1,662 (62.\$) | 2,379 (89.3) | 2,109 (79.1) | 1,182 (44.3) | 2,267 (85.1) |

| C. KD signs by age groups |  |  |  |  |  |
| --- | --- | --- | --- | --- | --- |
|  |  |  | Number (%) |  |  |
| Peng<sup>23</sup> |  |  | < 1 yr | 1-5 yr | >5 yr |
|  |  |  | n=62 | n=286 | n=50 |
|  |  | BCGitis | 27 (43.5) | 18 (63.0) | 0 (0) |
|  |  | Conjunctivitis | 56 (90.3) | 242 (84.6) | 41 (82.0) |
|  |  | Extremities<br>changes | 22 (35.5) | 130 (45.5) | 16 (32.0) |
|  |  | Lymphadenopathy | 18 (29) | 183 (64) | 32 (54) |
|  |  | Oral lesions | 54 (87.1) | 234 (81.8) | 38 (76.0) |
|  |  | Rash | 34 (54.8) | 186 (64.3) | 26 (52) |

| Ruan<sup>24</sup> |  |  | < 6 months | 6 months-5 yr | >5 years |
| --- | --- | --- | --- | --- | --- |
|  |  |  | n=49 | n=1,160 | n=165 |
|  |  | BCGitis | 9 (18.4) | 39 (3.4) | 0 (0) |
|  |  | Conjunctivitis | 46 (93.9) | 1,112 (95.9) | 152 (94.4) |
|  |  | Extremities changes | 29 (59.2) | 922 (79.5) | 110 (68.3) |
|  |  | Lymphadenopathy | 7 (14.3) | 364 (31.4) | 99 (61.5) |
|  |  | Oral lesions | 36 (73.5) | 1,043 (89.9) | 143 (88.8) |
|  |  | Rash | 43 (87.8) | 966 (83.3) | 118 (73.3) |

D. Prevalence of signs (%) in complete and incomplete KD
| Complete KD |  |  | Incomplete KD |  |  |
| --- | --- | --- | --- | --- | --- |
|  | Seo<sup>6</sup> |  |  | Seo<sup>6</sup> | Bae<sup>28</sup> |
| BCGitis | 98.1 |  | BCGitis | 100 | 70.8 |
| Lymphadenopathy | 16 |  | Lymphadenopathy | 34 | 20.8 |
| Conjunctivitis | 100 |  | Conjunctivitis | 75.9 | NA |
| Extremities | 92.6 |  | Extremities | 20.7 | NA |
| Lips, mucosa | 98.1 |  | Lips, mucosa | 79.3 | 29.1 |
| Rash | 52 |  | Rash | 65.5 | 50 |

#### 3.5.2 Comparison with other KD signs

Seven studies compared BCGitis with other KD signs (Table 3B). Its prevalence (3.5– 70.8%) was lower than most other signs, but occasionally exceeded lymphadenopathy, particularly in younger children (<1 year: 43.5% vs 29% and <6 months: 18.4% vs 14.3%) (Table 3C). Comparing complete with incomplete KD forms (Table 3D), one study by Seo demonstrated that BCGitis was less frequent in complete than in incomplete KD, while the second study by Bae only analysed incomplete forms. However, in both studies, the prevalence of BCGitis exceeded that of lymphadenopathy and rash.

### 3.6 BCG vaccination

#### 3.6.1 Interval between BCGitis and BCG vaccination

Limited data were available. One study reported intervals of 30–806 days. Children with BCGitis had significantly shorter vaccination-to-symptom intervals (hazard ratio 0.995, 95% CI 0.993–0.997, p<0.001) (Supplementary file: Table 2A).

#### 3.6.2 BCGitis prevalence by BCG vaccine strain

Eleven studies (73%) included patients vaccinated with Tokyo 172-1 (n=5), Pasteur 1173- P2 (n=3), Danish SSI-1331 (n=1), and Chinese substrains of Danish (n=2). The prevalence of BCGitis was 42.7–43.6% for Tokyo, Pasteur, and Danish strains, but much lower for Chinese substrains (5.2%) (Supplementary file: Table 2B).

### 3.7 Response to therapy by BCGitis status

Three studies (n=17,742) assessed the response to intravenous immunoglobulin (IVIG) administration. No significant differences were found between children with and without BCGitis (59.0% vs 57.2%) (Table 4).

**Table 4.**
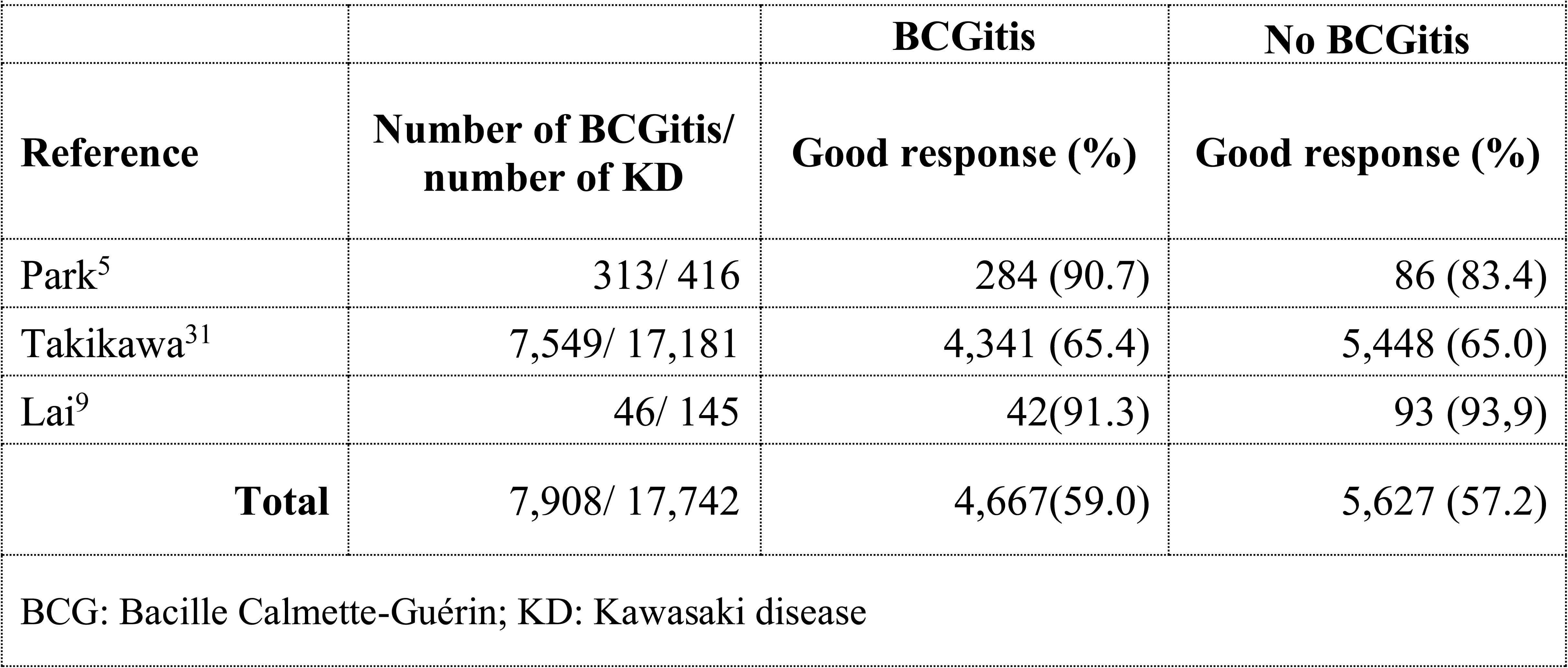
BCGitis in KD and good response to intravenous immunoglobulins.

### 3.8 Coronary artery abnormalities (CAA) by BCGitis status

This was reported in four studies (n=17,920). Three studies reported separately coronary dilatation and aneurysm, with only one report merging both. Two reports showed lower rates of coronary dilatation in children with BCGitis (11.6% vs 13.8%; 20.0% vs 22.0%; 4.3% vs 7.1%), while one showed slightly higher prevalence without BCGitis (11.8% vs 7.7%). Aneurysms were absent in two reports, with only one study showing marginally lower rates without BCGitis (3.0 vs 4.3%) (Supplementary file: Table 3A). In seven studies, the overall CAA prevalence was 12.3% for children vaccinated with the Tokyo strain and 11.8% for those with the Pasteur strain (Supplementary file: Table 3B).

### 3.9 Synthesis of results

From 15 studies from five Asian countries (Japan, Korea, Taiwan, China, Singapore), including 21,880 children with KD, the overall prevalence of BCGitis was 40.1%. Younger children consistently showed a higher prevalence. BCGitis was associated with lower CAA rates (11.8% vs 13.8%) but showed no significant association with KD type, vaccine strain, or treatment response

## 4. DISCUSSION

### 4.1 Summary of evidence

This scoping review provides the first comprehensive synthesis of information about BCGitis in Kawasaki disease (KD). It was performed on data from 15 studies conducted in five Asian countries, encompassing > 20,000 children. Despite heterogeneity among these studies in their design, the overall prevalence of BCGitis was approximately 40%, supporting earlier findings.^15,16^ This further reinforces its position as a reliable diagnostic marker instead of a coincidental discovery. The prevalence did not differ by BCG vaccine strain, suggesting that host response, rather than vaccine composition, is the primary determinant.

There was a significant inverse association between BCGitis prevalence and age, with younger children being the most affected and having a higher risk of incomplete KD and cardiovascular problems. ^7,8,16,17^ This finding, correlating with a shorter interval since receiving the vaccine in early infancy, may likely reflect greater immunological activity at the BCG site in infancy. ^8,18–20^ Although a male predominance was noted, contrary to earlier investigations, this difference could not be explained. ^8,11^ In agreement with other research, the prevalence of BCGitis did not vary between full and incomplete KD. ^8,21^ This emphasises its potential utility as a diagnostic tool in both forms and reinforces its application as an auxiliary criterion to mitigate diagnostic delays.

The review corroborates several prior studies where coronary artery anomalies were more prevalent in children without BCGitis. ^5,7,8,20^ It has been suggested that this is because BCGitis is recognised earlier than other signs in KD, leading to earlier treatment.

Alternatively, it may also delineate a KD phenotype with increased cutaneous involvement and diminished coronary vasculitis. The lack of correlation between BCGitis and IVIG resistance reinforces the notion that it signifies a distinct immunophenotype rather than general disease severity.

### 4.2 Strengths

This review focused exclusively on KD in BCG-vaccinated children, ensuring internal validity because unvaccinated children are incapable of developing BCGitis. All the reviewed studies systematically examined BCG vaccination sites. The substantial cumulative sample size (>20,000 patients) and geographic variety across various Asian nations augment the generalizability of these results among vaccinated populations. The age-related pattern of BCGitis occurrence and its correlation with reduced CAA rates were constant across various study designs and healthcare systems, hence reinforcing the conclusions.

### 4.3 Limitations

There are a few limitations that need to be recognised. Most of the reviewed studies were retrospective, making them susceptible to selection and detection bias, especially if doctors were aware of the KD diagnosis before assessing BCG sites. Some case reports have indicated that BCGitis can appear by day 3 or 4 of KD, occasionally preceding other signs. ^22^ The studies included in this review, however, did not report on this specific timing relative to other signs of KD. ^18,19,22^ This, therefore, hinders the evaluation of its efficacy as an early diagnostic indicator in KD. In addition, individual patient-level data were unavailable, thus preventing multivariate analysis to adjust for confounders such as treatment timing, sex, or vaccination age. Standardised definitions of BCGitis and CAA were sometimes lacking, with several investigations amalgamating coronary dilation and aneurysms. All included studies were conducted in Asian countries with universal BCG vaccination. The findings, therefore, limit their relevance to other contexts. Differences in timing, strain, and administration technique were rarely detailed, though they may influence BCGitis occurrence. While publication bias may exist, the inclusion of research with relatively low prevalence indicates that negative findings were also documented. Lastly, diagnostic accuracy metrics such as sensitivity and specificity could not be calculated due to the lack of an appropriate control group (BCGitis in non-BCG-vaccinated children).

Other constraints must also be acknowledged when interpreting these findings. The decision to include prevalence calculations is not common in scoping reviews, though this approach was justified because the primary outcome was homogeneous and summary estimates were clinically important. In line with scoping review methodology, we did not evaluate the quality of the included studies; hence, we may not have identified all pertinent methodological considerations.

### 4.4 Recommendations for research

Future studies should establish standardised definitions and assessment protocols for BCGitis. They should include a consensus on optimal examination timing and documentation methods. Prospective studies collecting individual-level data are needed to evaluate their diagnostic and prognostic value while controlling for confounding variables. Longitudinal studies could clarify onset, evolution, and resolution patterns of BCGitis.

Multicenter studies in diverse geographic and ethnic populations are essential to ascertain generalizability. The notably lower prevalence reported in the two studies from China warrants further investigation into strain-related or population or methodological factors.^23,24^ Clinical trials should determine if systematic evaluation of BCGitis improves diagnostic accuracy, reduces treatment delays, and lowers CAA risk. Due to its ease and lack of expense, routine evaluation of BCG sites may be a useful addition to KD diagnostic algorithms, particularly in populations that have received the BCG vaccine.

### 4.5 Conclusions

Conclusive recommendations await well-designed prospective studies. However, existing evidence suggests an important role for BCGitis as a useful supportive evidence for KD, especially in younger children and incomplete KD cases. Healthcare practitioners in BCG-vaccinated populations should routinely evaluate BCG vaccination sites in febrile children suspected of KD without incurring additional costs or resources. However, increasing population mobility leads to the inclusion in these countries of a pool of children vaccinated with BCG in their original country. Therefore, systematic assessment of BCGitis remains relevant and may aid earlier KD diagnosis even in countries without universal BCG vaccination.

## Supporting information

Suopplements 1 to 3

## Data Availability

All data produced in the present work are contained in the manuscript

## ETHICAL APPROVAL

None required

## DATA SHARING

All the data supporting the findings of this study are available from the cited references.

## AUTHOR STATEMENTS

HN conceptualised and designed the review, had primary responsibility for protocol development, literature search, screening of titles and abstracts, retrieving and reading the eligible studies, extracting and analysing the data, writing the first draft of the manuscript and approving the final version. AS participated in the screening of titles and abstracts, reading the eligible studies, extraction of the data, contributed to the writing of the manuscript and approved the final version

