## Supplement 4 for "Bacillus Calmette-Guérin scar reactivation (BCGitis) in Kawasaki disease. A Scoping Review"

### Supplementary File 4- Search Strategies

#### PubMed

*Core search (MeSH + text words):*

(
"Kawasaki Disease"[Mesh] OR "Kawasaki disease"[tiab] OR "Kawasaki's disease"[tiab]
OR "mucocutaneous lymph node syndrome"[tiab] OR MCLS[tiab]
OR "atypical Kawasaki"[tiab] OR "incomplete Kawasaki"[tiab] OR KD[tiab]
)
AND (
"Bacillus Calmette-Guérin Vaccine"[Mesh]
OR "Bacille Calmette-Guerin"[tiab] OR "Bacillus Calmette-Guerin"[tiab] OR BCG[tiab]
OR "BCG scar"[tiab] OR BCGitis[tiab] OR "BCG reactivation"[tiab] OR "BCG scar reactivation"[tiab]
OR "BCG vaccination site"[tiab] OR "BCG inoculation site"[tiab] OR "BCG site"[tiab] OR "BCG-site"[tiab]
OR "BCG site erythema"[tiab] OR "erythema at BCG site"[tiab] OR "scar erythema"[tiab]
OR "injection site erythema"[tiab] OR "inoculation site erythema"[tiab]
)

("Case Reports"[Publication Type] OR "Observational Study"[Publication Type] OR "Cohort Studies"[Mesh] OR "Cross-Sectional Studies"[Mesh])

#### Scopus

*Core search (TITLE-ABS-KEY):*

TITLE-ABS-KEY(
"Kawasaki disease" OR "Kawasaki's disease" OR "mucocutaneous lymph node syndrome" OR MCLS
OR "atypical Kawasaki" OR "incomplete Kawasaki" OR KD
)
AND TITLE-ABS-KEY(
"Bacille Calmette-Guerin" OR "Bacillus Calmette-Guerin" OR BCG OR "BCG scar" OR BCGitis
OR "BCG reactivation" OR "BCG scar reactivation" OR "BCG vaccination site" OR "BCG inoculation site"
OR "BCG site erythema" OR "erythema at BCG site" OR "scar erythema" OR "injection site erythema" OR "inoculation site erythema"
)

AND TITLE-ABS-KEY(cohort OR "cross-sectional" OR observational)

#### EMBASE

('BCGitis' OR 'bcg scar reactivation' OR ‘Bacille Calmette-Guerin’ OR ‘BCG’ OR ‘BCG’ scar’ OR ‘BCG vaccination site’ OR ‘BCG inoculation site’ OR ‘BCG site erythema’ OR ‘erythema at BCG site’ OR ‘injection site erythema’ OR ‘inoculation site erythema’) AND ('kawasaki disease' OR 'kawasaki disease' OR ‘mucocutaneous lymph node syndrome’ OR ‘MCLS’ OR ‘incomplete Kawasaki’ OR ‘atypical Kawasaki)

#### Cochrane Library (Advanced Search)

Core search using MeSH and keywords:

#1 MeSH descriptor: [Kawasaki Disease] explode all trees
#2 ("Kawasaki disease" OR "Kawasaki's disease" OR "mucocutaneous lymph node syndrome" OR MCLS OR "incomplete Kawasaki" OR "atypical Kawasaki" OR KD):ti,ab,kw
#3 MeSH descriptor: [Bacillus Calmette-Guérin Vaccine] explode all trees

#4 ("Bacille Calmette-Guerin" OR "Bacillus Calmette-Guerin" OR BCG OR "BCG scar" OR BCGitis OR "BCG reactivation" OR "BCG scar reactivation"
OR "BCG vaccination site" OR "BCG inoculation site" OR "BCG site erythema" OR "erythema at BCG site" OR "injection site erythema" OR "inoculation site erythema"):ti,ab,kw
#5 #1 OR #2
#6 #3 OR #4
#7 #5 AND #6

#10 ("cohort OR "cross-sectional" OR observational):ti,ab,kw
#11 #7 AND #10

**Web of Science (Core Collection)**

TS=(
"Kawasaki disease" OR "Kawasaki's disease" OR "mucocutaneous lymph node syndrome" OR MCLS OR "incomplete Kawasaki" OR "atypical Kawasaki" OR KD
)
AND TS=(
"Bacille Calmette-Guerin" OR "Bacillus Calmette-Guerin" OR BCG OR "BCG scar" OR BCGitis OR "BCG reactivation" OR "BCG scar reactivation"
OR "BCG vaccination site" OR "BCG inoculation site" OR "BCG site erythema" OR "erythema at BCG site" OR "injection site erythema" OR "inoculation site erythema"
)

AND TS=(" cohort OR "cross-sectional" OR observational)
