## Supplement 5 for "Bacillus Calmette-Guérin scar reactivation (BCGitis) in Kawasaki disease. A Scoping Review"

### Supplementary File 5. Data Collection Form

Primary author's name: ____________________

Year of publication: _____________________

Study period: from ___________ to _____________

Country: ______________________________

Study type:

Cohort observational ☐ Case-control ☐ Other type: ________________

Study setting: Health care setting ☐ other: ______________________

BCG vaccination status: Specifically confirmed ☐ Universal policy ☐ Not mentioned ☐

BCG vaccine strain: ______________________________

Age of participants: from ____ to _____

or mean ± SD: ☐ median (IQR): ☐ Not mentioned ☐

BCGitis presence: Yes ☐ No ☐ NA ☐

Number of children with KD: ______________

Number of children with BCGitis: __________

BCGitis sex group comparison: Yes ☐ No ☐ NA ☐

BCGitis age group comparison: Yes ☐ No ☐ NA ☐

BCGitis and KD type: Complete ☐ Incomplete ☐ Not mentioned ☐

BCGitis comparison with other KD signs: Yes ☐ No ☐

BCGitis timing compared to other KD signs: Yes ☐ No ☐

BCGitis timing after BCG vaccination: Yes ☐ No ☐ NA ☐

BCGitis and coronary abnormalities:

Coronary dilatation: Yes ☐ No ☐ NA ☐

Coronary aneurysm: Yes ☐ No ☐ NA ☐

BCGitis and good response to IVIG: Yes ☐ No ☐ NA ☐
