## Supplementary material for "Bacillus Calmette-Guérin scar reactivation (BCGitis) in Kawasaki disease. A Scoping Review": Suopplements 1 to 3

### Supplementary file: Table 1. BCGtis association with age and sex in KD

#### A. Prevalence of BCGitis by age group

| References | Age group comparison | Number of BCGitis/ number of KD | BCGitis prevalence (%) |
|---|---|---|---|
| Ruan[24] | < 6 m | 9/49 | 18.4 |
| Loh[1] | < 1 yr | 30/ 99 | 30.3 |
| Peng[23] | | 27/ 62 | 43.5 |
| Kang[27] | | 16 / 20 | 80 |
| Teng[26] | | 50/ 109 | 45 |
| Loh[1] | > 1 yr | 89/ 180 | 49.4 |
| Teng[26] | | 30/ 242 | 16.1 |
| Park[5] | < 18 m | 313/ 418 | 74.9 |
| Park[5] | > 18 m | 130/ 180 | 72.2 |
| Lai[9] | < 2 yr | 46/ 100 | 46 |
| Takikawa[31] | | 7,026/ 11,372 | 61.8 |
| Kim[30] | | 62/ 114 | 54.4 |
| Lai[9] | > 2 yr | 0/ 46 | 0 |
| Takikawa[31] | | 527/ 5,809 | 9.1 |
| Kim[30] | | 15/ 185 | 8.1 |
| Ruan[24] | 6 m- 5 yr | 39/1,160 | 3.4 |
| Peng[23] | 1-5 yr | 18/ 286 | 63 |
| Peng[23] | > 5 yr | 0/ 50 | 0 |
| Ruan[24] | | 0/161 | 0 |

| **B. Prevalence of BCGitis by sex** | | | | |
|---|---|---|---|---|
| **References** | **Males with BCGitis/ KD** | **Prevalence in males (%)** | **Females with BCGitis/KD** | **Prevalence in females (%)** |
| Takikawa[31] | 4,578/ 9,993 | 45.8 | 2,971/ 7,188 | 41.3 |
| Park[5] | 197/ 263 | 74.9 | 116/ 153 | 75.8 |
| Loh[1] | 76/ 172 | 44.2 | 43/ 107 | 40.2 |
| Lai[9] | 31/ 90 | 34.4 | 15/ 55 | 27.3 |
| **Total** | 4,882/ 10,518 | 46.4 | 3,145/ 7,503 | 41.9 |

KD: Kawasaki Disease; BCG: Bacille Calmette-Guérin; BCGitis/ KD= number with BCGitis/ number of KD

### Supplementary file: Table 2. BCG vaccination in KD and BCGitis

#### A. Timing interval of BCGitis since vaccination

| | | Number of children | |
|---|---|---|---|
| **Reference** | **Days after vaccination** | **BCGitis** | **No BCGitis** |
| Araki[29] | <30 | 0 | 4 |
| | 31-60 | 8 | 0 |
| | 61-806 | 71 | 21 |
| | ≧ 807 | 0 | 69 |

The interval from vaccination to symptom onset was identified as being most closely positively associated with BCGitis (hazard ratio, 0.995; $p < 0.001$; 95% confidence interval,0.993–0.997).

#### B. BCGitis prevalence by the BCG vaccination strain used

| References | Country | BCG strain | Number of BCGitis/ number of KD | BCGitis prevalence (%) |
|---|---|---|---|---|
| Loh[1] | Singapore | Danish SSI 1331 | 119/ 279 | 42.70 |
| Seo[6] | Korea | Pasteur 1173-P2 | 83/ 490 | 16.90 |
| Park[5] | Korea | Pasteur 1173-P2 | 313/ 416 | 75.20 |
| Kang[27] | Korea | Pasteur 1173-P2 | 24 /64 | 37.50 |
| Lin[11] | Taiwan | Tokyo 172-1 | 85/ 280 | 30.40 |
| Lai[9] | Taiwan | Tokyo 172-1 | 46/ 145 | 31.70 |
| Araki[29] | Japan | Tokyo 172-1 | 83/ 177 | 46.90 |
| Yokota[25] | Japan | Tokyo 172-1 | 13/ 18 | 72.20 |
| Takikawa[31] | Japan | Tokyo 172-1 | 7,549/ 17,181 | 43.90 |
| Ruan[24] | China | Chinese substrain (Beijing D1), Shanghai D2PB302 (derived from Danish strain 823 | 48/ 1370 | 3.5 |
| Peng[23] | China | Chinese substrain (Beijing D1), Shanghai D2PB302 (derived from Danish strain 823 | 45/ 398 | 11.2 |

| References | Country | BCG strain | Number of BCGitis/ number of KD | BCGitis prevalence (%) |
|---|---|---|---|---|
| Total | | Danish SSI | 119/ 279 | 42.70 |
| | | Pasteur 1173-P2 | 420/ 970 | 43.30 |
| | | Tokyo 172-1 | 7,776/ 17,801 | 43.60 |
| | | Chinese substrain (Beijing D1), Shanghai D2PB302 (derived from Danish strain 823 | 93/ 1768 | 5.2 |

BCG: Bacille Calmette-Guérin; KD: Kawasaki disease

### Supplementary file: Table 3. BCGitis and coronary artery abnormalities in KD

#### A. Prevalence of coronary artery abnormalities (CAA) by BCGitis status

| | | | BCGitis present | | BCGitis absent | |
|---|---|---|---|---|---|---|
| **Reference** | **Number of BCGitis/ number of KD** | **CAA type** | **Number of CAA** | **Prevalence (%)** | **Number of CAA** | **Prevalence (%)** |
| Takikawa[31] | 7,549/ 17,181 | Dilatation or aneurysm | 873/ 7,459 | 11.6 | 1,337/ 9,632 | 13.8 |
| Park[5] | 313/ 416 | Dilatation | 37/ 313 | 11.8 | 8/ 103 | 7.7 |
| | | Aneurysm | 0/313 | 0.0 | 0/313 | 0.0 |
| Araki[29] | 83/ 178 | Dilatation | 17/ 83 | 20.0 | 21/ 94 | 22.0 |
| | | Aneurysm | 0/83 | 0.0 | 0/94 | 0.0 |
| Lai[9] | 46/145 | Dilatation | 2 /46 | 4.3 | 7 / 99 | 7.1 |
| | | Aneurysm | 2/46 | 4.3 | 3/99 | 3.0 |

#### B. Prevalence of CAA in children with BCGitis by the BCG vaccine strain

| References | BCG strain | Number of KD with BCGitis | Number with CAA | CAA prevalence (%) |
|---|---|---|---|---|
| Takikawa[31] | Tokyo 172-1 | 7459 | 875 | 12 |
| Park[5] | Pasteur 1173-P2 | 313 | 37 | 11.8 |
| Araki[29] | Tokyo 172-1 | 83 | 17 | 20.5 |
| Lai[9] | Tokyo 172-1 | 46 | 2 | 4.3 |
| Lin[11] | Tokyo 172-1 | 44 | 24 | 54.5 |
| Kang[27] | Tokyo 172-1 | 24 | 7 | 29.1 |
| Yokota[25] | Tokyo 172-1 | 13 | 2 | 15.4 |
| **Total** | Pasteur 1173-P2 | 313 | 37 | 11.8 |
| | Tokyo 172-1 | 7759 | 948 | 12.3 |

BCG: Bacille Calmette-Guérin; KD: Kawasaki disease; CAA: coronary artery abnormalities
